# Hypomethylation of miR-17-92 cluster in lupus T cells and no significant role for genetic factors in the lupus-associated DNA methylation signature

**DOI:** 10.1101/2022.02.21.22271293

**Authors:** Patrick Coit, Xiavan Roopnarinesingh, Lourdes Ortiz-Fernandez, Kathleen Maksimowicz-McKinnon, Emily E. Lewis, Joan T. Merrill, W. Joseph McCune, Jonathan D. Wren, Amr H. Sawalha

**Affiliations:** Division of Rheumatology, Department of Pediatrics, University of Pittsburgh, Pittsburgh, Pennsylvania, USA; Graduate Program in Immunology, University of Michigan, Ann Arbor, Michigan, USA; Graduate Program, Department of Biochemistry and Molecular Biology, The University of Oklahoma Health Sciences Center, Oklahoma City, Oklahoma, USA; Arthritis and Clinical Immunology Program, Oklahoma Medical Research Foundation, Oklahoma City, Oklahoma, USA; Division of Rheumatology, Henry Ford Health System, Detroit, Michigan, USA; Division of Rheumatology, Department of Internal Medicine, University of Michigan, Ann Arbor, Michigan, USA; Department of Biochemistry and Molecular Biology, The University of Oklahoma Health Sciences Center, Oklahoma City, Oklahoma, USA; Division of Rheumatology and Clinical Immunology, Department of Medicine, University of Pittsburgh, Pennsylvania, USA; Lupus Center of Excellence, University of Pittsburgh School of Medicine, Pittsburgh, Pennsylvania, USA; Department of Immunology, University of Pittsburgh, Pittsburgh, Pennsylvania, USA

## Abstract

**Objectives:** Epigenetic dysregulation plays an important role in the pathogenesis of lupus, a systemic autoimmune disease characterized by autoantibody production. Lupus T cells demonstrate aberrant DNA methylation patterns dominated by hypomethylation of interferon-regulated genes. The objective of this study was to identify additional disease-associated DNA methylation changes in naïve CD4+ T cells from an extended cohort of lupus patients and determine the genetic contribution to epigenetic changes characteristic of lupus.

**Methods:** Genome-wide DNA methylation was assessed in naïve CD4+ T cells isolated from a cohort of 74 lupus patients and 74 age-, sex-, and race-matched healthy controls. We applied a trend deviation analysis approach, comparing methylation data in our cohort to methylation data from over 16,500 samples to characterize lupus-associated DNA methylation patterns. Methylation quantitative trait loci (meQTL) analysis was used to determine genetic contribution to lupus-associated DNA methylation changes.

**Results:** In addition to the previously reported epigenetic signature in interferon-regulated genes, we observed hypomethylation of the promoter regions of microRNA (miRNA) genes in the miR-17-92 cluster in lupus patients. Members of this miRNA cluster play an important role in regulating T cell proliferation and differentiation. Expression of two miRNAs within this cluster, miR-19b1 and miR-18a, showed a significant positive correlation with disease activity in lupus patients. meQTL were identified by integrating genome-wide DNA methylation profiles with genotyping data in lupus patients and controls. Patient meQTL show overlap with genetic risk loci for lupus. However, less than 1% of differentially methylated CpG sites in lupus patients were associated with an meQTL, suggesting minimal genetic contribution to lupus-associated epigenotypes.

**Conclusion:** The lupus defining epigenetic signature, characterized by robust hypomethylation of interferon-regulated genes, does not appear to be determined by genetic factors. Hypomethylation of the miR-17-92 cluster that plays an important role in T cell activation is a novel epigenetic locus for lupus.

## Introduction

Systemic lupus erythematosus (lupus or SLE) is a heterogeneous autoimmune disease of incompletely understood etiology. The disease is characterized by a loss of immunotolerance and the development of autoantibodies against nuclear antigens. Severe manifestations of lupus have significant impact on quality of life and can lead to organ damage and mortality in affected patients, particularly among patients of non-European genetic ancestry [1, 2]. Genetic risk contributes to the development of lupus, but the estimated heritability of lupus is ~30% [3–5]. Indeed, monozygotic twin studies in lupus suggest a substantial non-genetic contribution to the etiology of lupus [6]. Environmental exposures across the lifespan that can directly impact epigenetic regulation and cellular function are suggested to be involved in the pathogenesis of lupus [7, 8].

DNA methylation is an epigenetic mechanism that regulates gene expression through the enzyme-mediated addition of a methyl group to the cytosine bases in the genome. DNA methylation is heritable across cell generations and can promote gene silencing, making it an important component in regulating the plasticity of immune cell identity and function [9]. Early work demonstrated that adoptive transfer of CD4+ T cells treated *ex vivo* with DNA methyltransferase (DNMT) inhibitors was sufficient to cause lupus-like disease in mice [10] mimicking the DNA methylation inhibition in patients with drug-induced lupus [11]. Since then, other studies have observed that CD4+ T cells of lupus patients show a distinct shift in global DNA methylation compared to healthy individuals, potentially in part due to defective MEK/ERK signaling, suppressing DNMT1 activity in CD4+ T cells, and leading to hypomethylation and overexpression of costimulatory genes [12–16].

We have previously observed a robust hypomethylation signature in interferon-regulated genes defining lupus patients [17, 18]. Our initial findings in CD4+ T cells were subsequently confirmed and expanded to other cell types by our group and others [19–21]. In CD4+ T cells, we observed hypomethylation in interferon-regulated genes at the naïve CD4+ T cell stage, preceding transcriptional activity. This epigenetic “poising” or “priming” of interferon-regulated genes was independent of disease activity [18]. The genetic contribution to this lupus-associated epigenotype is currently unknown.

Methylation quantitative trait loci (meQTL) are genetic polymorphisms that are associated with the methylation state of CpG sites either through direct nucleotide change within the CpG dinucleotide or intermediary mechanisms. Prior studies of lupus patients show an enrichment of meQTL associated with type I interferon genes, genetic risk loci, and specific clinical manifestations in whole blood and neutrophils [22–24]. Furthermore, our previous work suggests that meQTL might at least in part explain differences in DNA methylation between African-American and European-American lupus patients [22].

Herein, we evaluated genome-wide DNA methylation data in naïve CD4+ T cells from a large cohort of lupus patients compared to matched healthy controls. We integrated DNA methylation and genotyping data to better understand the influence of genetic factors upon the DNA methylation changes observed in lupus.

## Methods

### Study Participants and Demographics

74 female lupus patients and 74 female healthy age (± 5 years), race, and sex-matched controls were recruited as previously described [25, 26] (**Supplementary Table 1**). All patients fulfilled the American College of Rheumatology (ACR) classification criteria for SLE [27]. Institutional review boards at our participating institutions approved this study. All participants signed a written informed consent prior to participation.

### Sample collection, DNA isolation, and data generation

Genomic DNA samples for this study were collected from naïve CD4+ T cells as previously described [18]. Briefly, magnetic beads and negative selection was used to isolate naïve CD4+ T cells from whole blood samples collected from lupus patients and controls. Genomic DNA was directly isolated from collected cells and bisulfite converted using the EZ DNA Methylation Kit (Zymo Research, Irvine, CA, USA). The Illumina HumanMethylation450 BeadChip (Illumina, San Diego, CA, USA) was used to measure DNA methylation levels at over 485,000 methylation sites across the genome.

### Epigenome-wide association study

Epigenome-wide association study (EWAS) for identifying associations between specific CpG sites and disease status was performed using GLINT [28, 29]. Covariates for age, race, and technical batch were included for the analysis prior to other preprocessing. No outliers beyond four standard deviations were detected in the first two components of PCA space, all 148 samples were included in the analysis. Reference-less cell type composition correction was performed using *ReFACTor*, with six components used in the downstream analysis to account for any cell-type heterogeneity in the samples. An additional covariate was included to account for effects of genetic admixture using the EPISTRUCTURE algorithm included in GLINT. Cell-type composition covariate components generated by *ReFACTor* were included at this step to reduce bias from potential cell-type heterogeneity, and polymorphic CpG sites were excluded from this step and the EWAS. Using the initial age, race, and technical batch covariates, along six *ReFACTor* components and one *EPISTRUCTURE* component, logistic regression for disease status was performed across all CpG sites, excluding the polymorphic and unreliable cross-reactive probes previously described in the literature, as well as CpG sites with low variance (standard deviation <0.01) [30, 31].

### Differential DNA methylation analysis of gene promoters

Raw .idat files were used to generate methylation beta value profiles across all samples using GenomeStudio (Illumina, San Diego, CA, USA) after background subtraction and normalizing to internal control probes. Missing probe values were imputed using *sklearn*.*impute*.*KNNImputer*, and *ComBat* was used to correct for batch effects associated with technician sample preparation [32–34]. Ensembl gene loci for hg19 were downloaded using *PyEnsembl* [35]. For each gene, loci for 1500 base pairs upstream of the transcription start site [36] to the TSS were mapped to the overlapping CpG probes using *PyBedtools*, and the mean of the associated probes for each gene was used as the representative methylation value for the resulting 20,437 mapped genes [37]. Differential methylation analysis comparing patients and controls was performed on the mean TSS1500 methylation using *limma*, and false discovery rate adjustment using the Benjamini-Hochberg method was used to correct P-values for multiple testing. Gene Ontology Enrichment for Biological Process terms was performed on the differentially methylated gene list using *Enrichr* with the mapped promoter gene list used as the background [38, 39].

### Trend deviation analysis

DNA methylation data derived using the Illumina 450k methylation array from 23,415 samples were downloaded from Gene Expression Omnibus (GEO) [40]. To reduce batch effects, samples from experiments with less than 50 samples were omitted, and the resulting samples were quantile normalized [41]. A matrix of pairwise Pearson’s correlation values for DNA methylation levels was computed across TSS1500 gene promoters in 16,541 samples across 37 tissues to create a multi-tissue correlation network (**Supplementary Figure 1**). The differentially methylated genes in lupus naïve CD4+ T cells were clustered by their correlation in global signature created from the GEO data. Hierarchical clustering was performed using *Scipy*’s hierarchical linkage. KEGG enrichment analysis was performed using *Enrichr* [42], and P-values were reported after false-discovery rate adjustment.

The goal of a trend deviation analysis is to detect correlation patterns among differentially methylated genes in large DNA methylation datasets. A correlation in methylation among a set of differentially methylated genes between patients and controls suggests a trend is being observed, reinforcing the significance and robustness of the differential DNA methylation detected between patients and controls.

### Genotyping

Genomic DNA isolated from naïve CD4+ T cells was used as input for the Infinium Global Screening Array-24 v2.0 (Illumina, San Diego, CA, USA). Single nucleotide polymorphisms (SNPs) with a genotyping call rate < 98%, minor allele frequencies (MAF) < 5%, and deviating from Hardy-Weinberg equilibrium (HWE; P-value < 1E-3) were filtered out. Samples were removed if they had a genotyping call rate < 95%. Sex chromosomes were not analyzed. About 100,000 independent SNPs were pruned and used to perform principal component analysis (PCA) with *Eigensoft* (v.6.1.4) software [43]. Genotyping data were analyzed using *PLINK* (v.1.9) [44]. Genotype profiles were generated for n = 63 patients and n = 68 controls.

### Methylation Quantitative Trait Loci (meQTL) Analysis

Raw .idat files were used to generate methylation profiles using *minfi* (v.1.32.0) [45, 46] and to check median intensity values and reported sex in the R statistical computing environment (v.3.6.3) [47]. Probes with less than three beads and zero intensity values across all samples were removed using the *DNAmArray* package (v.0.1.1) [48]. Background signal and dye bias were corrected, followed by normalization of signal intensities using functional normalization in the *preprocessFunnorm*.*DNAmArray* function [48, 49] using the first three principal component values calculated from signal intensities of control probes present on all array spots to correct for technical variation. Any probe with a detection

P-value < 0.01 or returned signal intensities in fewer than 98% of samples was removed. This resulted in a total of 476,767 probes used for further analysis. Signal intensities were then converted to M-values with a maximum bound of ±16. M-values were used for meQTL analysis and converted to beta values (0-100% methylation scale) using *minfi* for reporting.

We removed any probe for meeting any of the following technical criteria: A unique probe sequence of less than 30bp, mapping to multiple sites in the genome, polymorphisms that cause a color channel switching in type I probes, inconsistencies in specified reporter color channel and extension base, mapping to the Y chromosome, and/or having a polymorphism within 5bp of the 3’ end of the probe with a minor allele frequency > 1% with the exception of CpG-SNPs with C>T polymorphisms which were retained [50]. Batch correction for chip ID was performed using the *ComBat* function in the *sva* (v.3.34.0) package [51]. After technical filtering, there were a total of 421,214 probes used for meQTL analysis.

We implemented a mixed correspondence analysis with the *PCAmixdata* package (v.3.1) [52] to calculate eigenvalues using patient medication data for prednisone, hydroxychloroquine, azathioprine, mycophenolate mofetil, and cyclophosphamide. The top four components accounted for a cumulative 89.3% of variability in the medication data. Each component value was used as an independent variable in regression analysis to adjust for medication usage across individuals. MeQTL association analysis was performed in patients and controls separately using methylation M-value profiles and corresponding sample genotypes. Age, the top four medication components, and top ten genotype principal components were included as covariates to build a linear model for detecting meQTL using *MatrixEQTL* (v.2.3) [53]. *Cis*-meQTL were defined as CpG sites with methylation values associated with a SNP within a conservative 1000bp of the CpG dinucleotide. We used a Benjamini-Hochberg FDR-adjusted P-value cutoff of < 0.05 as a threshold for significant associations. The above EWAS results were compared with the meQTL results to determine overlap of lupus-associated differentially methylated CpG sites and those CpG sites in an meQTL.

### Gene Set Enrichment Analysis (GSEA)

ToppGene Suite was used for functional gene ontology enrichment analysis [54] of Molecular Function and Biological Process Gene Ontologies and KEGG Pathways in meQTL loci. P-values were derived using a hypergeometric probability mass function, and a Benjamini-Hochberg FDR–adjusted P-value cutoff of < 0.05 was used as a threshold of significance. A minimum membership of 3 genes and maximum of 2000 genes in each term was used as a threshold for inclusion. IFN-regulated genes were identified using the set of genes associated with the CpG site in each meQTL as input using Interferome (v.2.01) [55]. The type I interferon response genes were defined as genes with an expression fold change of 1.5 or greater between type I interferon-treated and untreated samples using gene expression datasets from all available CD4+ T cell experiments in the Interferome database.

### MicroRNA Expression Microarray

MicroRNA (miRNA) expression was measured in naïve CD4+ T cells from a subset of lupus patients and healthy matched controls (n = 16). Cells were immediately lysed with TRIzol Reagent (ThermoFisher Scientific, NY, USA) followed by storage at −80C. Total RNA was isolated using the Direct-zol RNA MiniPrep Kit (Zymo Research, CA, USA) following the manufacturer’s directions. The Affymetrix miRNA 4.1 Array Strip (Affymetrix, CA, USA) was used to measure expression of over 2,000 premature and 2,500 mature human miRNA sequences. RNA sequences were polyadenylated and ligated to a biotin-labeled oligomer using the FlashTag Biotin HSR RNA Labeling Kit (Affymetrix, CA, USA). Biotin-labeled sequences were hybridized to array probes and washed then stained with streptavidin-PE. The Affymetrix Expression Console & Transcriptome Analysis Console 2.0 software (Affymetrix, CA, USA) was used to analyze biotin/streptavidin-PE fluorescence measurements. All samples passed signal intensity, polyadenylation, and ligation quality controls. Signal intensities were background adjusted and normalized. Log2-transformed expression values for each probeset was calculated using a robust multi-array average model (23). The Pearson *r* correlation coefficient for median expression values of probes for miR-17, miR-18a, miR-19a, miR-19b1, and miR-20a and patient SLEDAI score were calculated using GraphPad Prism (v9.3.0) (GraphPad Software, CA, USA).

## Results

### Differential methylation of gene promoters in naïve CD4+ T cells isolated from lupus patients

A comparison of DNA methylation profiles from circulating naïve CD4+ T cells isolated from 74 lupus patients and 74 age, sex and race matched healthy controls revealed a total of 2,627 CpGs out of 334,337 total CpG sites included in the EWAS with a significant differential methylation. Significant hypomethylation in interferon-regulated genes was observed, consistent with previous reports (**Supplementary Table 2**). Average promoter methylation for each gene was calculated by including all CpG sites on the array within 1500bp of the associated gene’s transcription start site (TSS). A total of 51 genes showed a significant difference in average promoter methylation between lupus patients and controls (17 hypomethylated and 34 hypermethylated in patients compared to controls) (**Table 1**) (**Figure 1**). Biological Process Gene Ontology enrichment analysis of differentially methylated promoter regions did not show significant enrichment compared to the background of all gene promoters after adjusting for multiple testing (**Supplementary Table 3**).

**Table 1.** Genes with differentially methylated promoter regions in naive CD4+ T cells of lupus patients compared to healthy controls.

| **Gene** | **$\Delta\beta$** | **$-\log_{10}$ (FDR-adjusted P-value)** | **t-statistic** |
| --- | --- | --- | --- |
| *IFI44L* | -0.177 | infinity | -10.757 |
| *DTX3L* | -0.130 | infinity | -11.566 |
| *BST2* | -0.089 | 11.323 | -9.285 |
| *RABGAP1L* | -0.088 | 9.165 | -8.421 |
| *BCL2L14* | -0.086 | 5.520 | -6.908 |
| *MIR19B1* | -0.059 | 3.169 | -5.846 |
| *IFI44* | -0.059 | 2.057 | -5.304 |
| *MIR20A* | -0.055 | 3.088 | -5.807 |
| *MIR17* | -0.054 | 6.882 | -7.487 |
| *MIR18A* | -0.051 | 6.537 | -7.342 |
| *MIR19A* | -0.049 | 4.771 | -6.579 |
| *IKZF4* | -0.048 | 3.289 | -5.902 |
| *MX1* | -0.046 | 10.624 | -9.004 |
| *TRIM34* | -0.045 | 2.184 | -5.367 |
| *ODF3B* | -0.034 | 1.712 | -5.128 |
| *GNG2* | -0.033 | 2.138 | -5.344 |
| *FAM177B* | -0.025 | 1.897 | -5.223 |
| *MZF1* | 0.008 | 1.493 | 5.014 |
| *SSBP4* | 0.015 | 1.344 | 4.934 |
| *ATP6V0D1* | 0.018 | 2.594 | 5.569 |
| *DCUN1D1* | 0.025 | 2.068 | 5.309 |
| *C14orf93* | 0.025 | 1.922 | 5.236 |
| *TIPARP* | 0.026 | 2.069 | 5.310 |
| *LMBRD1* | 0.027 | 2.211 | 5.381 |
| *HAVCR2* | 0.027 | 2.574 | 5.560 |
| *KIAA1949* | 0.030 | 3.158 | 5.841 |
| *GPD2* | 0.032 | 1.953 | 5.251 |
| *CNTF* | 0.033 | 1.705 | 5.124 |
| *CD47* | 0.034 | 4.259 | 6.350 |
| *ARHGAP9* | 0.036 | 3.339 | 5.926 |
| *IL27RA* | 0.036 | 1.367 | 4.946 |
| *RAP1A* | 0.036 | 2.573 | 5.559 |
| *LAMA3* | 0.037 | 1.445 | 4.988 |
| *ABI3* | 0.037 | 1.436 | 4.983 |
| *FAM102A* | 0.038 | 3.161 | 5.842 |
| *CXCR5* | 0.039 | 1.439 | 4.985 |
| *DPEP2* | 0.040 | 1.889 | 5.219 |
| *DYRK2* | 0.041 | 3.924 | 6.197 |
| *TMEM71* | 0.044 | 2.757 | 5.649 |
| *ADORA2A* | 0.046 | 2.234 | 5.392 |
| *SEPT9* | 0.047 | 2.036 | 5.293 |
| *PSMB4* | 0.052 | 2.935 | 5.734 |
| *TOM1* | 0.055 | 5.415 | 6.862 |
| *PRIC285* | 0.057 | 9.934 | 8.729 |
| *LTB* | 0.062 | 2.036 | 5.293 |
| *MIR1205* | 0.067 | 1.698 | 5.121 |
| *ACER3* | 0.073 | 2.612 | 5.578 |
| *BCL9L* | 0.079 | 4.034 | 6.248 |
| *MDS2* | 0.080 | 3.149 | 5.836 |
| *SNORA5B* | 0.083 | 1.712 | 5.128 |
| *PTPRCAP* | 0.091 | 3.620 | 6.057 |

FDR correction was performed using the Benjamini-Hochberg method with an FDR-adjusted P value threshold of < 0.05. $\Delta\beta$ : methylation difference in median methylation value of CpG sites within 1500bp upstream of the associated gene's transcription start site (TSS1500) between lupus patients and healthy controls.

**Figure 1.**
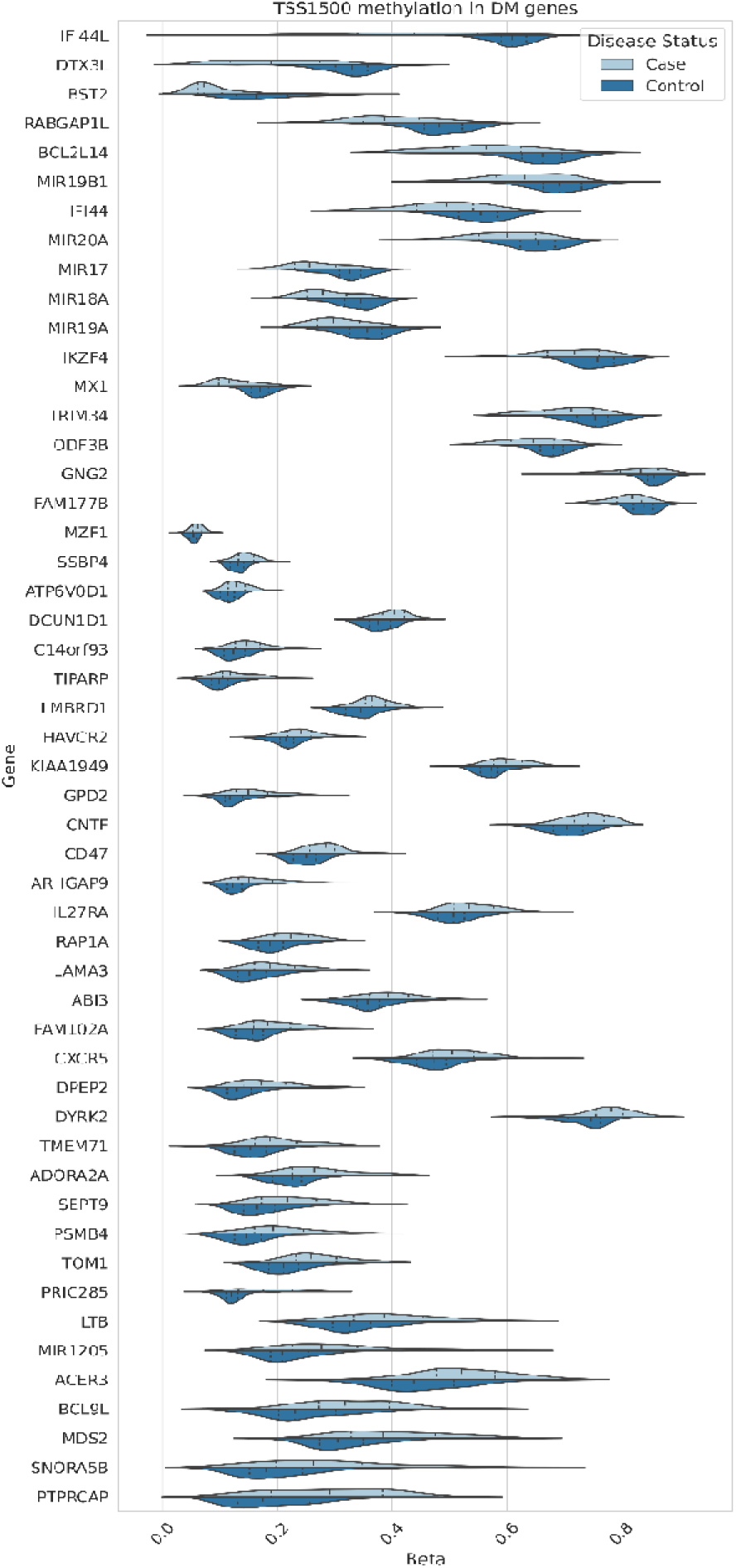
Distribution of average CpG methylation levels within 1500bp of the TSS for the respective genes differentially methylated in naïve CD4+ T cells of lupus patients compared to healthy controls.

The pairwise correlation of the 51 gene promoters identified above was calculated across a collection of 16,541 samples from 37 tissues available in GEO. Hierarchical clustering of correlations showed that 21 out of the 51 gene promoters were highly correlated. KEGG Pathway enrichment analysis showed a significant enrichment for three pathways among the 21 correlated gene promoters: “microRNAs in cancer” (P-value = 3.86E-04), “cytokine-cytokine receptor interaction” (P-value = 4.34E-02), and “rheumatoid arthritis” (P-value = 4.34E-02) (**Table 2**) (**Figure 2**). The microRNAs in cancer” pathway included genes encoding miR-17, miR-18a, miR-19a, miR-19b1, and miR-20a. Four of seven CpG sites used to calculate the average promoter methylation (TS1500) in this locus showed a significant reduction in median methylation in lupus patients compared to healthy controls (**Figure 3A**). These sites: cg17799287 (Δβ= −5.5%; P-value = 2.05E-03), cg07641807 (Δβ = −4.4%; P-value = 1.71E-02), cg23665802 (Δβ= −5.8%; P-value = 1.19E-02), and cg02297838 (Δβ= −4.9%; P-value = 3.48E-02) were all hypomethylated in lupus patients compared to healthy controls and overlapped with enhancers and regions flanking TSSs in peripheral naïve CD4+ T cells using data collected from the Epigenome Roadmap [56] and visualization using the WashU Epigenome Browser [57]. We examined expression levels of the microRNAs included in the “microRNAs in cancer” pathway (miR-17, miR-18a, miR-19a, miR-19b1, and miR-20a) in naïve CD4+ T cells of a subset of our lupus patients (n = 16) and healthy matched controls (n = 16). We did not observe a difference in expression between patients and control. However, two miRNAs, miR-18a-5p and miR-19b1-5p, showed a significant positive correlation (hsa-miR-18a-5a P-value = 0.038 & hsa-miR-19b1-5p P-value = 0.042) between median expression level and SLEDAI scores in lupus patients (**Figure 3B**) (**Supplementary Table 4**).

**Table 2.** KEGG Pathway gene enrichment of 21 gene promoters highly correlated with each other in multi-tissue DNA methylation data constructed from 16,541 samples available through Gene Expression Omnibus.

| **Pathway**<br>**(KEGG_2019_Human)** | **P-value** | **FDR-**<br>**adjusted**<br>**P-value** | **Odds**<br>**Ratio** | **Genes** |
| --- | --- | --- | --- | --- |
| MicroRNAs in cancer | 1.21E-05 | 0.00039 | 20.92 | *MIR19B1;MIR20A;MIR17;MIR18A;MIR19.* |
| Cytokine-cytokine<br>receptor interaction | 0.0034 | 0.043 | 11.28 | *CNTF;CXCR5;LTB* |
| Rheumatoid arthritis | 0.0041 | 0.043 | 23.52 | *LTB;ATP6V0D1* |

**Figure 2.**
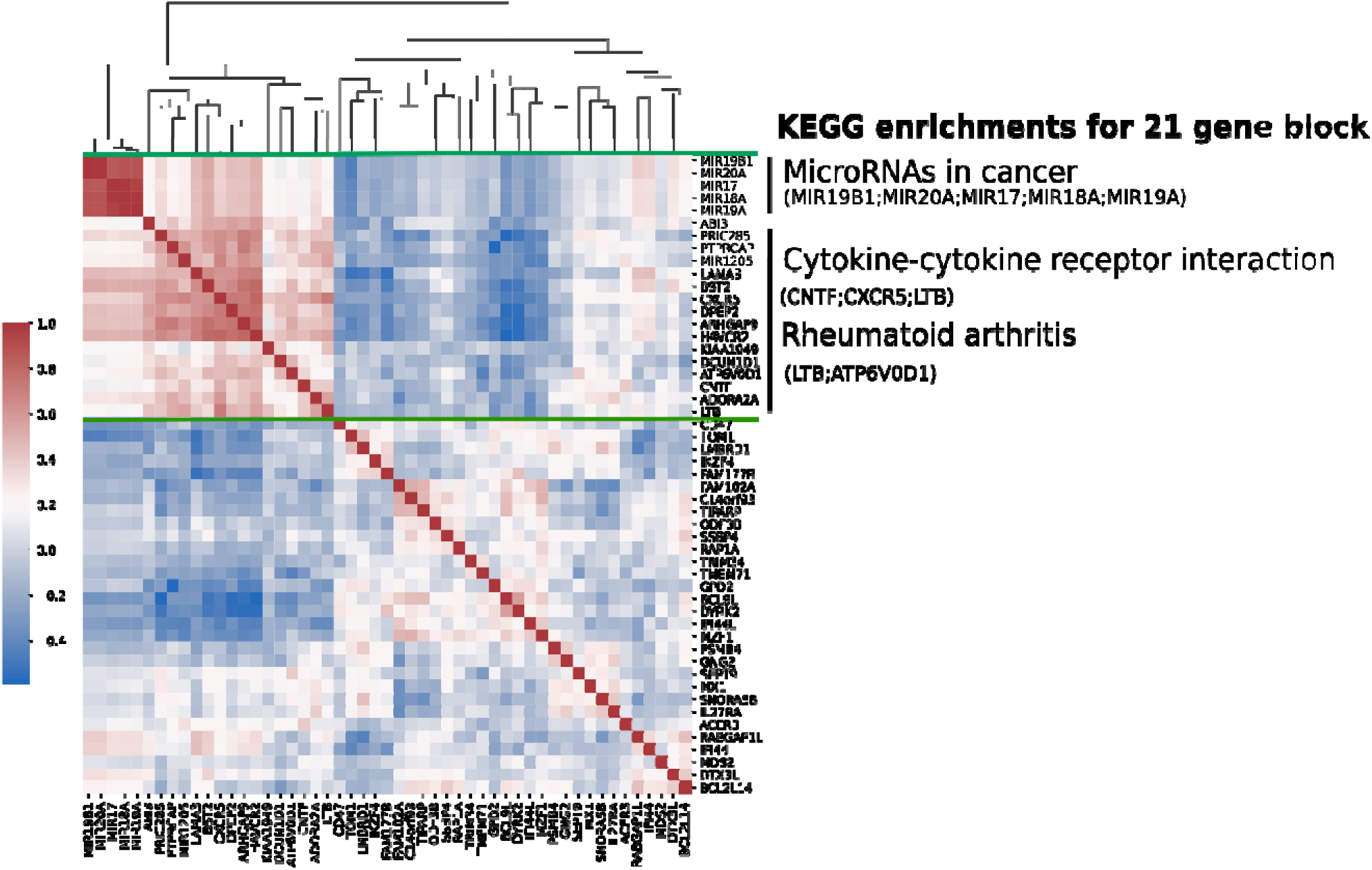
Heatmap of hierarchical clustering of pairwise Pearson correlation coefficient values of 51 differentially methylated gene promoters (TSS1500) in global tissue signature derived from 16,541 samples. Range from +1 (red) to −1 (blue), represent a greater to lower correlation in global tissue, respectively. KEGG pathways are significantly enriched (FDR-adjusted P-value < 0.05) in a block of 21 genes (green bars).

**Figure 3.**
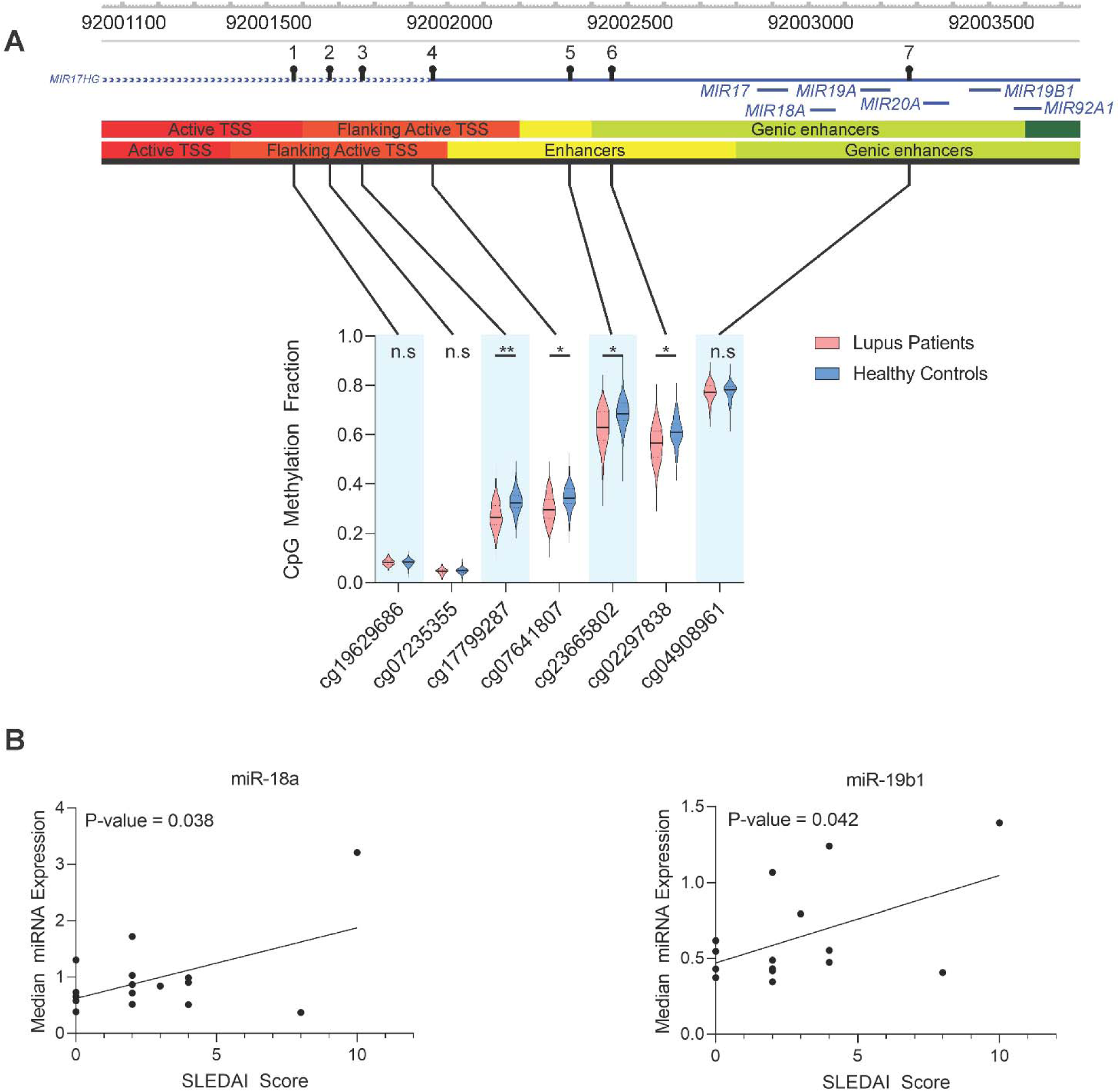
**(A)** Violin plots of the seven CG probes in lupus patients and healthy controls used to calculate the average promoter methylation (TSS1500) for the miR-17-92 cluster. The solid black bar represents the median value and the dashed lines the first and third quartiles. Genomic visualization and annotation are from WashU Epigenome Browser using AuxillaryHMM tracks from peripheral naïve CD4+ T cell tissues (E038 and E039, top and bottom tracks, respectively). For P-values: n.s. = not significant, * = P < 0.05, ** = P < 0.01. **(B)** Correlation of median miRNA expression in naïve CD4+ T cells of a subset (n = 16) of lupus patients with SLEDAI score. Hsa-miR-18a-5p and hsa-miR-19b1-5p had a Pearson correlation (r) of 0.52 (P-value = 0.038) and 0.51 (P-value = 0.042), respectively.

### Naïve CD4+ T cell methylation quantitative trait loci (meQTL) in lupus patients

Global genotype profiles were generated in a subset of patients and controls and compared with global DNA methylation profiles to identify CpG sites with allele-specific methylation associations. There was no significant difference in the average age between the patient (n = 63) and control (n = 68) subsets (patient average age = 41.6; patient age SD = 12.8; control average age = 40.8; control age SD = 12.5; t-test statistic = 0.3811; two-tailed P-value = 0.7038). Allele-specific DNA methylation associations were measured as meQTL where the CpG site was within 1000bp of the measured SNP separately in patients and controls. After adjusting for age, genetic background, and medication use in patients, we identified 5,785 meQTL present in the naïve CD4+ T cells of lupus patients with an FDR-adjusted P-value < 0.05 (**Supplementary Table 5**). These meQTLs represented 4,649 (80.4%) unique CpG sites and 4,120 (71.2%) unique polymorphisms. Of the 4,791 meQTL with a CpG-associated gene annotation, 2,356 (49.2%) were unique.

A linear model adjusting for age and genetic background was fit to controls separately. We identified a total of 7,331 meQTL with an FDR-adjusted P-value < 0.05 in controls (**Supplementary Table 6**). These meQTLs represented 5,885 (80.3%) unique CpG sites and 5,138 (70.1%) unique polymorphisms. Of the 6,061 meQTL with a CpG-associated gene annotation, 2,846 (47.0%) were unique.

We compared meQTL in lupus patients and healthy controls with the 2,627 CpG sites differentially methylated between the two groups. Of these, we identified 17 (0.6%) and 34 (1.3%) unique CpG sites with a significant change in DNA methylation in lupus patients and healthy controls, respectively (**Figure 4A and B**). We examined the overlap of meQTL in lupus patients and healthy controls and identified a total of 3,957 meQTL (68.4% of lupus patient meQTL and 54.0% of healthy control meQTL) shared between both patients and control meQTL sets (**Supplementary Table 7**). This shared set of meQTL contained 8 (0.3% of differentially methylated CpG sites) unique CpG sites that we identified as differentially methylated between lupus patients and controls (**Figure 4C**).

**Figure 4.**
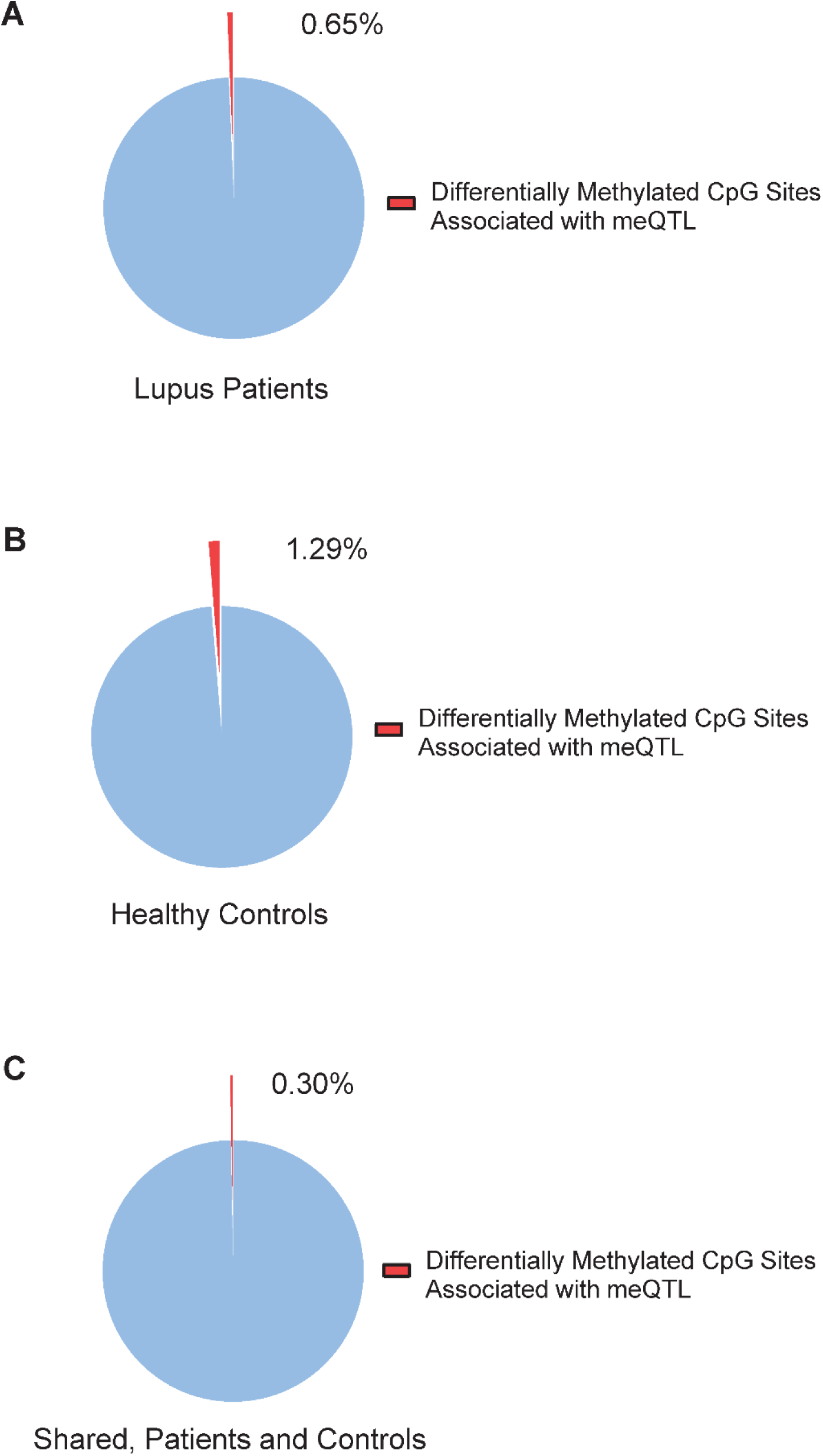
Proportion of differentially methylated CpG sites in naïve CD4+ T cells of lupus patients compared to healthy controls associated with an meQTL in **(A)** lupus patients, **(B)** healthy controls, and **(C)** the subset of meQTL shared between lupus patients and healthy controls.

Gene set enrichment analysis was performed using genes associated with CpG sites in our meQTL shared between patients and controls. GSEA revealed multiple ontologies and pathways for cell adhesion (“cell-cell adhesion”; P-value = 1.04E-12, “biological adhesion”; P-value = 6.80E-12, and “cell adhesion”; P-value = 8.25E-12, “Cell adhesion molecules (CAMs)”; P-value = 2.25E-06), transporter associated with antigen processing (TAP) proteins and antigen presentation (“TAP binding”; P-values = 1.59E-7, “peptide antigen binding”; P-value = 4.40E-5), immune disorder pathways (“Type I diabetes mellitus; P-value = 1.92E-8, and “Graft-versus-host disease”; P-value = 4.38E-7) (**Supplementary Table 8**).

There were 1,828 meQTL detected only in lupus patients but not in controls. These were enriched in gene ontologies and pathways related to tissue growth and development (“animal organ morphogenesis”; P-value = 8.44E-10, “urogenital system development”, P-value = 1.05E-07) and gene silencing (“negative regulation of gene silencing by miRNA”; P-value = 2.54E-6, “negative regulation of posttranscriptional gene silencing”; P-value = 5.41E-6) (**Supplementary Table 9**).

We compared our list of meQTL in lupus patients to previously identified lupus susceptibility loci from genome-wide association studies[4, 58–61] and found 41 meQTL contained CpG site-associated genes that overlapped with 20 unique risk loci genes (**Supplementary Table 10**). This included interferon regulatory factor genes *IRF5* and *IRF7*. We examined the overlap of our meQTL-associated genes identified in lupus patient naïve CD4+ T cells and genes that respond to type I interferon treatment in CD4+ T cells to better understand the association between patient genetics and type I interferon-response gene methylation differences in lupus. A total of 101 unique type I interferon-response genes were meQTL loci in our data (**Supplementary Table 11**).

## Discussion

We generated genome-wide DNA methylation data in naïve CD4+ T cells from a large cohort of lupus patients and matched healthy controls. Implementing an innovative trend deviation analysis, we identified a cluster of microRNAs (miRNAs) (miR-17, miR-18a, miR-19a, miR-19b1, miR-20a) among differentially methylated loci in lupus patients. Promoter methylation analysis revealed significant hypomethylation in this microRNA cluster in lupus patients compared to controls. Trend deviation analysis suggested a coordinated, disease-associated change in promoter methylation for these miRNAs. Indeed, the expression of miR-18a and miR-19b1 included within this cluster positively correlated with disease activity, as measured using SLEDAI score, in our lupus patients. MiRNAs play an important role in post-transcriptional gene regulation by targeting specific complementary gene transcripts for degradation [62]. Peripheral blood cells in lupus patients show altered expression of miRNAs [63]. Some deregulated miRNAs in lupus target DNA methyltransferase 1 (DNMT1), and as a result, contribute to altered DNA methylation patterns in lupus CD4+ T cells [64–66]. MiR-17, miR-18a, and miR-20a form the “miR-17 family” while miR-19a and miR-19b1 form the “miR-19 family”, which are grouped by sequence homology and encoded in a single polycistronic miRNA gene called the “miR-17-92 cluster”. This cluster has been well-studied as an oncogene and an immune regulator [67]. Average promoter methylation of miR-17, miR-18a, miR-19a, miR-19b1, and miR-20a was reduced by ~5% in lupus patients compared to controls, which has not been previously described in immune cells of lupus patients. Enterovirus 71 infection has been observed to suppress miR-17-92 cluster expression by increasing DNMT-mediated promoter methylation [68], and chemical inhibition of DNMT1 activity in bleomycin-induced lung fibrosis mouse model increases miR-17-92 cluster expression in lung fibroblasts [69]. This suggests that miR-17-92 cluster promoter methylation plays an important role in regulating the expression of its members.

MiR-17-92 cluster genes play a vital role in regulating T cell activities including proliferation and differentiation. Overexpression of miR-17-92 cluster genes promotes lymphoproliferative disease and autoimmunity in mice by targeting critical immunotolerance regulators Bim and PTEN [70]. Conditional knock out of miR-17-92 cluster in a murine model of chronic graft-versus-host disease (cGVHD) reduced disease-associated T cell infiltration and IgG deposition in the skin [71]. In cGVHD mice, miR-17-92 cluster expression in CD4+ T cells supports Th1, Th17, and Tfh cell differentiation. Loss of miR-17-92 cluster expression leads to a corresponding reduction in Tfh-dependent germinal center formation and plasma cell differentiation [71]. MiR-17, miR-18a, miR-19a, and miR-20a are overexpressed in splenic T cells of MRL/*lpr* mice [72]. Similarly, miR-17, miR-17a, miR-18a, miR-19a, miR-19b1, and miR-20a are overexpressed in circulating CD4+ T cells of lupus patients [73]. MiR-19b1 expression, specifically, has a significant positive correlation with disease activity as measured by SLEDAI score [73]. MiR-17 and miR-20 are downregulated in circulating PBMCs [74], B cells [75], as well as circulating free miRNAs [76] of lupus patients compared to healthy controls, suggesting tissue-specific and miR-specific expression patterns. Of the miR-17-92 cluster miRNAs identified as differentially methylated in our analysis, only miR-18a and miR-19b1 showed a significant positive correlation between median expression and disease activity in naïve CD4+ T cells of lupus patients, consistent with these prior observations. MiR-19b1 promotes proliferation of mature CD4+ T cells, Th1 differentiation and IFN-γ production, and suppresses inducible Treg differentiation [77]. MiR-18a expression increases rapidly early on in CD4+ T cell activation [78, 79], and suppresses Th17 cell differentiation through direct targeting of critical Th17 transcription factor transcripts including *Smad4, Hif1a*, and *Rora* in human CD4+ T cells *in vitro* and *in vivo* murine airway inflammation models [78]. We did not observe a difference in the expression of members in the miR-17-92 cluster between lupus patients and controls in naïve CD4+ T cells, likely because these miRNAs are upregulated upon T cell activation. Evidence for hypomethylation in lupus in naïve CD4+ T cells suggests epigenetic priming of this locus, similar what we previously observed in interferon-regulated gene loci in lupus [18]. Further study is needed to determine if altered DNA methylation at these miRNA promoter sites is associated with expression changes in miRNAs that play a role in T cell development and lupus pathogenesis and their potential use as a biomarker for monitoring disease activity.

We used analysis of meQTL to identify allele-specific DNA methylation associations across the genome of naïve CD4+ T cells from lupus patients and healthy controls. Our primary objective was to understand to what extent are DNA methylation changes associated with lupus (the lupus-defining epigenetic profile), explained by genetic factors. We found that < 1% of differentially methylated sites in lupus patients compared to healthy controls were associated with a *cis*-meQTL. This suggests that almost all of the DNA methylation alterations observed in lupus are not associated with local allelic differences in the genome, suggesting a greater contribution from non-genetic and possibly environmental factors to epigenetic dysregulation in lupus. A previous study of meQTL in whole blood of lupus patients found that a majority of meQTLs were shared between patients and controls [24]. We observed 68% of meQTL in lupus patients and 54% of meQTL in healthy controls were shared by both groups, supporting this observation. Gene ontology and pathway analysis of the meQTL-associated genes unique to lupus patients were enriched for KEGG pathways related to type I diabetes, viral myocarditis, and graft-versus-host disease all of which were primarily driven by the presence of meQTLs in HLA genes. Gene ontologies were related to the development of various tissues without an apparent relationship to disease pathogenesis. Of note, we did not observe meQTL effect involving the miR-17-92 cluster.

Our prior analysis of neutrophils from a cohort of lupus patients identified overlap in meQTL genes and lupus genetic risk loci [22]. MeQTL pairs including *ARID5B* (cg13344587-rs10821936), *HLA-DQB1* (cg13047157-rs9274477), and *IRF7* (cg16486109-GSA-rs1131665) were found in both neutrophils and naïve CD4+ T cells from lupus patients. Risk loci genes unique to naïve CD4+ T cell meQTLs included *CD80* (cg06300880-GSA-rs3915166), *TYK2* (cg06622468-rs280501), *IKBKE* (cg22577136-GSA-rs17020312), and *CTLA4* (cg05092371-GSA-rs16840252, cg05092371-rs4553808). Naïve CD4+ T cell-specific meQTL risk loci genes are all related to signal response and activation in CD4+ T cells compared to the more general DNA repair and type I interferon signaling seen in the shared meQTL risk loci genes. Disease-relevant meQTL show tissue-specific patterns which should be considered when teasing apart their potential impact.

In summary, we investigated genome-wide DNA methylation changes in naïve CD4+ T cells from an extended cohort of lupus patients and controls, and using a methylation trend deviation analysis method, we showed promoter hypomethylation of the miR-17-92 cluster that has a significant regulatory function in T cells growth, function, and differentiation. Combining genome-wide DNA methylation and genotyping data, we were able to determine genetic contribution to the lupus-defining epigenotype. Our data indicate that epigenetic changes characteristic of lupus are not under direct genetic influence. This suggests a more important role for non-genetic factors in the epigenetic dysregulation observed in lupus patients, including the robust demethylation of interferon-regulated genes.

## Data Availability

All data are included in supplementary material provided.

## Acknowledgements

This work was supported by the National Institute of Allergy and Infectious Diseases (NIAID) of the National Institutes of Health (NIH) grant number R01 AI097134 to Dr. Sawalha. Dr. Wren is supported by NIH grants number P30 AG050911 and P20 GM103636.

---

**Supplementary Figure 1.**
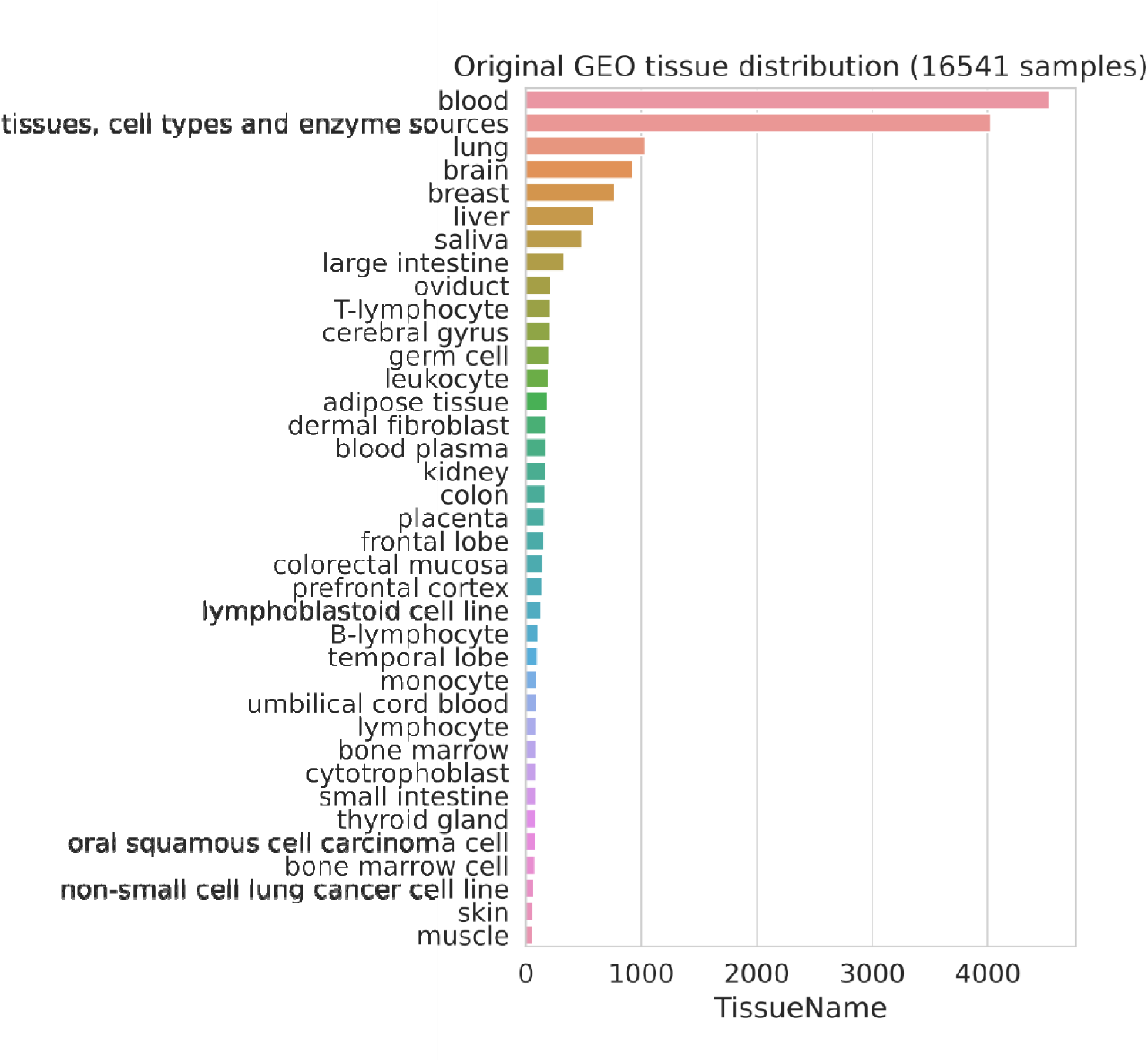
Sample distribution of 16,541Infinium HumanMethylation450 array samples across 37 tissues sourced from the Gene Expression Omnibus (GEO). These samples were used to develop a multi-tissue correlation network used for trend deviation analysis.

